# Predictive value of Serum CCL21 and CCL19 levels in heart failure patients : A prospective study

**DOI:** 10.1101/2023.10.10.23296854

**Authors:** Wenfei Zeng, Ling Li, Liman Wang, Biting Lin, Kailing Lin, Peng Yu, Huizhen Yu

## Abstract

**Background:** Chemokine C-C motif ligand (CCL)21 and CCL19 are well-recognized to associate with adverse events of cardiovascular disease, especially long-term prognosis. However, few studies have reported its correlation with heart failure.

**Purpose:** To investigate and compare the predictive value of CCL21 and CCL19 serum levels in patients with heart failure.

**Methods:** In this prospective, observational, single-center study, 221 patients with heart failure and 55 healthy controls were included. ROC curves were generated to analyze and compare the serum levels of CCL21 and CCL19 in predicting all-cause mortality and composite events. Cox regression and Kaplan-Meier survival analyses were performed to identify independent risk factors for prognosis. Pearson correlation was used to measure the correlation between creatinine and CCL21 / CCL19 levels.

**Results:** The study observed 108 events (30 deaths and 78 occurred composite endpoints) over a median follow-up of 494.5(231.5,950.0) days. CCL21 showed strong predictive value for both all-cause mortality (AUC were 0.694, *P*=0.001) and composite endpoints (AUC 0.661, *P*=0.006 and <0.001). while the combination of CCL21 and NT-proBNP further improved the predictive power, with AUC being 0.796 and 0.662 on all-cause mortality and composite endpoints, respectively (both *P*<0.001). K-M survival analysis revealed that patients with increased CCL21 and CCL19 exhibited higher all-cause mortality (both *P*<0.05). Meanwhile, higher incidence of composite endpoint events was also observed in patients with elevated CCL21 (*P*<0.05). Importantly, multivariate COX regression analysis demonstrated that smoking, higher level of CCL21 and ischemic heart disease were independent risk factors for all-cause mortality (*P*<0.001). Furthermore, diabetes and elevation of CCL21 were associated with an increased risk of composite endpoints (*P*<0.001). On the other hand, changes in CCL19 levels showed a graded association with worse renal function, resulting in a slight increasing trend in G3 and G4/5, (HR = 2.64, 95% CI= 2.26-2.66, vs. HR 2.67, 95% CI 2.57-2.91, overall interaction *P*<0.05), with higher concentrations in G3 and G4/5(HR = 3.67, 95% CI= 3.27-3.85, vs. HR 4.11, 95% CI 3.67-4.38; overall interaction *P*<0.05).

**Conclusions:** Serum concentrations of CCL21 and CCL19 were significantly elevated in heart failure patients. High level of CCL21 is an independent risk factor for the adverse events in heart failure and may complement the prediction of those events which are less affected by renal function.

## Introduction

Heart failure is a leading cause of death worldwide, imposing an increasing burden on health and economies in various countries, particularly developing ones [1]. However, heart failure patients have a poor long-term outlook, with mortality rates reaching 20% at one year and 50% at five years. Approximately 20-30% of discharged patients require readmission within 30 days, averaging 1.3 readmissions per year [1,2]. Early identification of risk factors in heart failure patients can positively impact prognosis and enable timely treatment. Although N-terminal pre-B-type natriuretic peptide (NT-proBNP) has become an essential biomarker for diagnosing heart failure [3], its value can be influenced by certain diseases or factors such as chronic renal failure, type 2 diabetes, and acute coronary syndrome [4]. Therefore, early identification and prevention of heart failure are crucial. Our research team has previously focused on independent risk factors for heart failure, as well as their association with disease progression and prognosis assessment markers [5]. CCL21 and CCL19 are termed homeostatic chemokine, that act through their shared receptor CCR7 to serve as immune surveillance and regulation of leukocyte movement and homing [6,7]. The immune system plays a critical role in the ventricle remodeling and contributes to both the inflammatory and repair phases [8]. Moreover, several studies have suggested that CCL21 and CCL19 are expressed in various non-lymphoid cells and may be closely associated with cardiovascular diseases such as coronary artery disease, acute coronary syndromes and pulmonary arterial hypertension in systemic sclerosis [9–11]. It has been showed that levels of inflammatory biomarkers are closely related to the clinical status, severity and prognosis of heart failure patients [12–14]. Some previous studies have found higher circulating concentrations of CCL21 and CCL19 in heart failure patients [15]. However, there have been no studies on the predictive value of serum CCL21 and CCL19 in heart failure patients. Therefore, this study aims to compare the serum levels of CCL21, CCL19 in heart failure patients and evaluate the prognostic value of these biomarkers for adverse events. The study hypothesized that CCL21 and CCL19 could provide additional insights into identifying prognostic risk factors in heart failure patients.

## Materials and methods

### Study Population

A total of 221 patients diagnosed with heart failure were consecutively enrolled at the Department of Cardiovascular Medicine, Fujian Provincial Hospital South Branch between January 2019 and October 2021. Among them, 51 were excluded for the following reasons: 30 due to acute myocardial infarction within 1 month, 6 due to severe infection, 2 due to malignancy, and 2 due to renal failure requiring dialysis. Additionally, there were 11 patients with incomplete data. Ultimately, a cohort of one hundred and seventy heart failure patients was included in this study (mean age = 74.8 years, male: female ratio =61.18%:38.82%). This study was conducted in accordance with the Helsinki Declaration and approved by the ethics committee of Fujian Provincial Hospital. Written informed consent was obtained from all participants. The collected data included general demographics, cardiac functional grading based on New York Classification of Cardiac Function (NYHA), laboratory tests results, echocardiograms index, comorbidities, etiology of heart failure and the medical regimen were recorded.

### Study Sample

Peripheral venous blood was collected the day after admission and frozen at −80°C after centrifugation. Serum levels of CCL21 and CCL19 were measured using enzyme-linked immunosorbent assay (ELISA) with detection limits of 0.5pg/ml and 5pg/ml, respectively (Boster, WuHan, HuBei, China). The plasma level of NT-proBNP was determined by Electrochemiluminescence immunoassay (ECLIA).

### Follow-up and Events

Regular outpatient visits or telephone calls were used for follow-up, which lasted 1.4 years (12 follow-up visits were missed, and 2 cases were lost to follow-up). Among the patients, 19 were classified as NYHA II, 75 as HYHA III, and 76 as NYHA IV. The outcomes measured included all-cause mortality and composite events such as heart failure readmission, acute myocardial infarction, stroke, heart failure readmission, acute myocardial infarction, and stroke. Medical records were reviewed to verify the information provided. All patients received appropriate medication based on their heart failure condition or any additional comorbidities they had (Figure1).

**Figure1.**
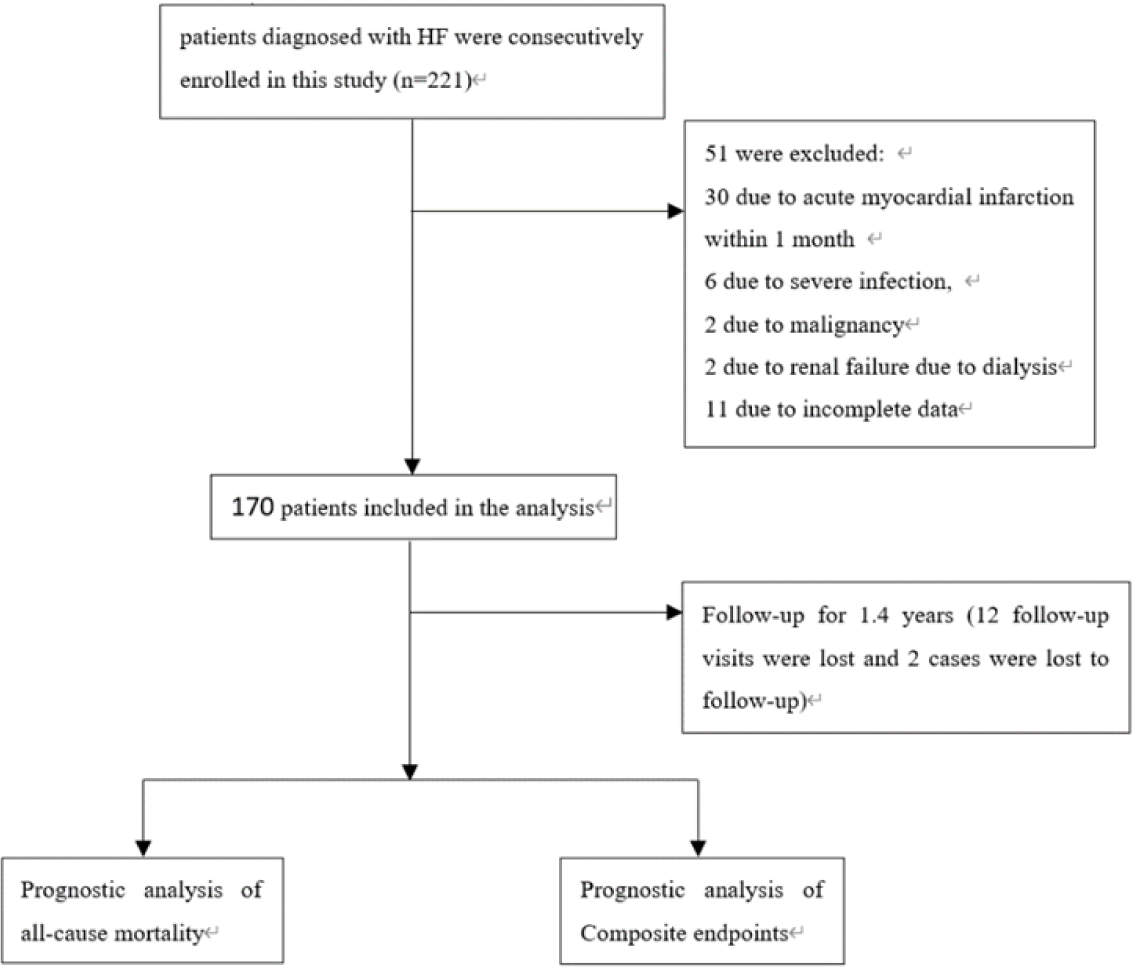
The design of the study

### Statistical Analyses

Continuous variables are presented as means±standard deviations or medians and interquartile ranges. Clinical data from participants with or without events were compared using Student’s t-test for normally distributed variables, Mann-Whitney U-test for skewed variables, and Chi-squared test for categorical parameters.

Levels of each biomarker with low or elevated values are classified based on the optimal cut-off value of the receiver-operating characteristic (ROC) curve analysis. The predictive ability of each variable was assessed by analyzing the area under the curve (AUC). Multivariate COX proportional hazard analyzes were conducted to identify independent risk factors for prognosis in heart failure patients. To investigate the time-to-event relationship for circulating biomarkers with opposite levels of CCL21, CCL19 and NT-proBNP, the K-M method was proposed and compared with a log-rank test. After adjusting for confounding factors, the hazard ratios (HRs) and 95% confidence intervals (CIs) were calculated for endpoints related to CCL21 and CCL19 levels. Kruskal-Wallis test was employed to compare biomarker levels among three groups of patients with different renal function. A two-sided *P* value<0.05 was considered statistically significant.

## Result

### Baseline Characteristics

Table 1 presents the clinical characteristics of study participants, categorized based on occurrence or nonoccurrence of events. Over a follow-up period of 1.4 years (with a median duration of 494.5 days), there were 30 (17.75%) deaths from any cause and 78 (45.88%) composite endpoints recorded. Age, systolic blood pressure (SBP), diastolic blood pressure (DBP), and body mass index (BMI) did not show significant differences between the groups with mortality/events and those without them. However, compared to the group without all-cause mortality, patients who experienced such mortality had higher proportions of men, smokers, CCL19 levels, CCL21 levels, NT-proBNP levels, and creatinine levels (P<0.05). Additionally, patients with composite endpoints exhibited higher values for CCL21 levels, NT-proBNP, NYHA Class III/IV, creatinine, hypertension, and diabetes severity than those without events. (*P* <0.05)

**Table 1.**
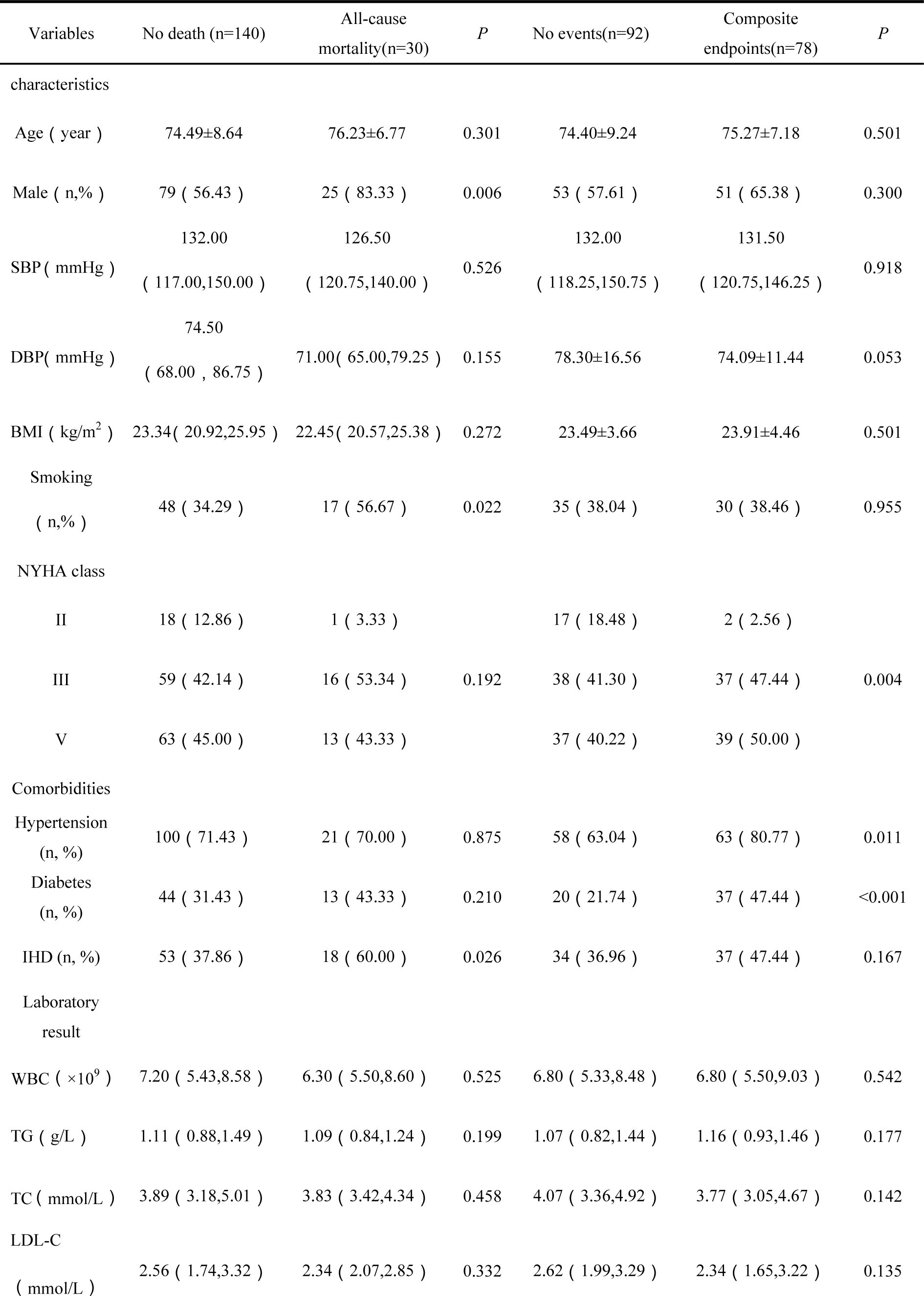
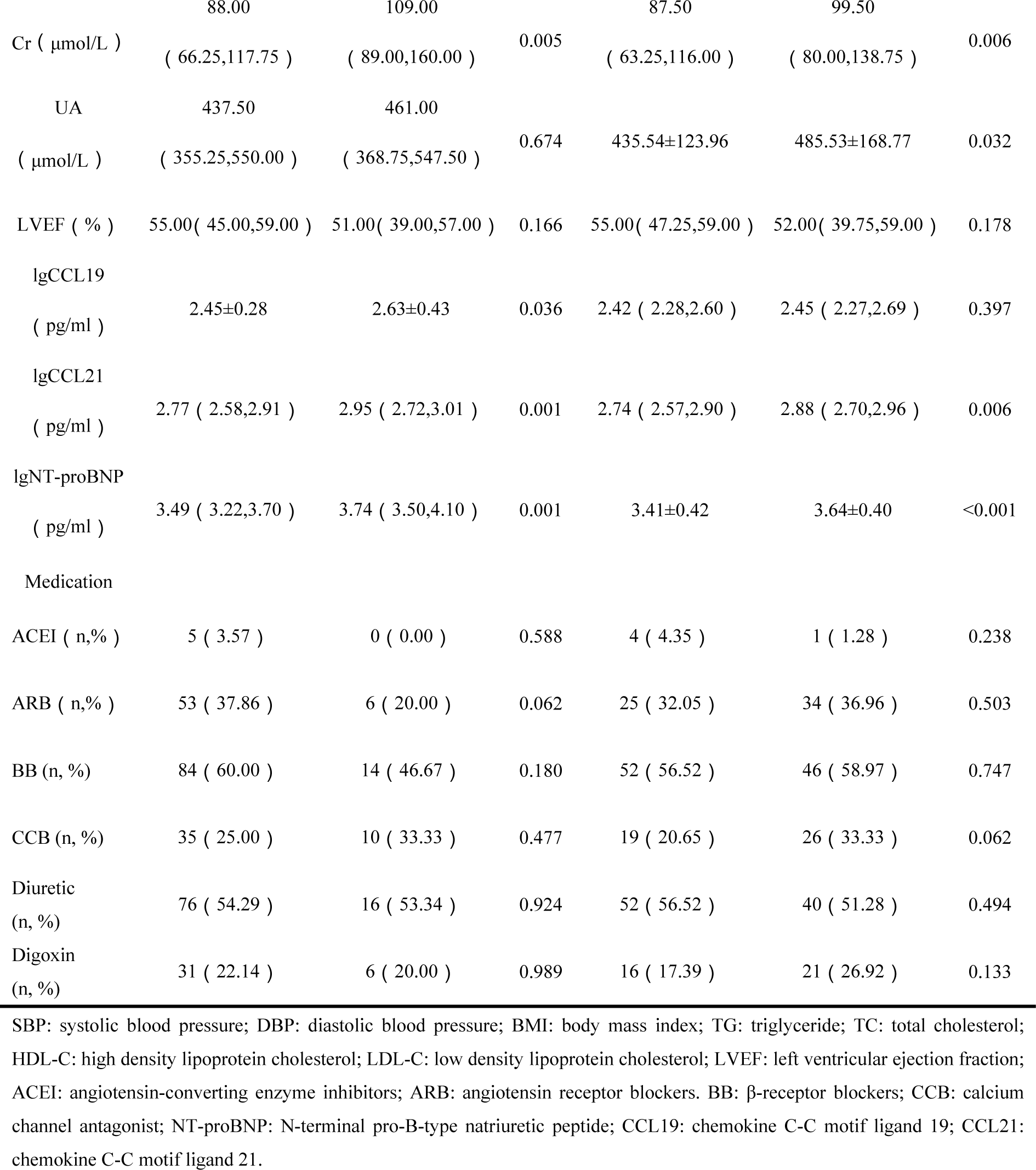
Baseline clinical characteristics of heart failure patients according to groups of all-cause mortality or composite endpoints occurred.

| Variables | No death (n=140) | All-cause mortality(n=30) | *P* | No events(n=92) | Composite endpoints(n=78) | *P* |
| --- | --- | --- | --- | --- | --- | --- |
| characteristics |  |  |  |  |  |  |
| Age ( year ) | 74.49±8.64 | 76.23±6.77 | 0.301 | 74.40±9.24 | 75.27±7.18 | 0.501 |
| Male ( n,% ) | 79 ( 56.43 ) | 25 ( 83.33 ) | 0.006 | 53 ( 57.61 ) | 51 ( 65.38 ) | 0.300 |
| SBP ( mmHg ) | 132.00<br>( 117.00,150.00 ) | 126.50<br>( 120.75,140.00 ) | 0.526 | 132.00<br>( 118.25,150.75 ) | 131.50<br>( 120.75,146.25 ) | 0.918 |
| DBP( mmHg ) | 74.50<br>( 68.00 , 86.75 ) | 71.00( 65.00,79.25 ) | 0.155 | 78.30±16.56 | 74.09±11.44 | 0.053 |
| BMI ( kg/m<sup>2</sup> ) | 23.34( 20.92,25.95 ) | 22.45( 20.57,25.38 ) | 0.272 | 23.49±3.66 | 23.91±4.46 | 0.501 |
| Smoking<br>( n,% ) | 48 ( 34.29 ) | 17 ( 56.67 ) | 0.022 | 35 ( 38.04 ) | 30 ( 38.46 ) | 0.955 |
| NYHA class |  |  |  |  |  |  |
| II | 18 ( 12.86 ) | 1 ( 3.33 ) |  | 17 ( 18.48 ) | 2 ( 2.56 ) |  |
| III | 59 ( 42.14 ) | 16 ( 53.34 ) | 0.192 | 38 ( 41.30 ) | 37 ( 47.44 ) | 0.004 |
| V | 63 ( 45.00 ) | 13 ( 43.33 ) |  | 37 ( 40.22 ) | 39 ( 50.00 ) |  |
| Comorbidities |  |  |  |  |  |  |
| Hypertension<br>(n, %) | 100 ( 71.43 ) | 21 ( 70.00 ) | 0.875 | 58 ( 63.04 ) | 63 ( 80.77 ) | 0.011 |
| Diabetes<br>(n, %) | 44 ( 31.43 ) | 13 ( 43.33 ) | 0.210 | 20 ( 21.74 ) | 37 ( 47.44 ) | <0.001 |
| IHD (n, %) | 53 ( 37.86 ) | 18 ( 60.00 ) | 0.026 | 34 ( 36.96 ) | 37 ( 47.44 ) | 0.167 |
| Laboratory result |  |  |  |  |  |  |
| WBC ( ×10<sup>9</sup> ) | 7.20 ( 5.43,8.58 ) | 6.30 ( 5.50,8.60 ) | 0.525 | 6.80 ( 5.33,8.48 ) | 6.80 ( 5.50,9.03 ) | 0.542 |
| TG ( g/L ) | 1.11 ( 0.88,1.49 ) | 1.09 ( 0.84,1.24 ) | 0.199 | 1.07 ( 0.82,1.44 ) | 1.16 ( 0.93,1.46 ) | 0.177 |
| TC ( mmol/L ) | 3.89 ( 3.18,5.01 ) | 3.83 ( 3.42,4.34 ) | 0.458 | 4.07 ( 3.36,4.92 ) | 3.77 ( 3.05,4.67 ) | 0.142 |
| LDL-C<br>( mmol/L ) | 2.56 ( 1.74,3.32 ) | 2.34 ( 2.07,2.85 ) | 0.332 | 2.62 ( 1.99,3.29 ) | 2.34 ( 1.65,3.22 ) | 0.135 |
| Cr ( $\mu\text{mol/L}$ ) | 88.00<br>( 66.25,117.75 ) | 109.00<br>( 89.00,160.00 ) | 0.005 | 87.50<br>( 63.25,116.00 ) | 99.50<br>( 80.00,138.75 ) | 0.006 |
| UA<br>( $\mu\text{mol/L}$ ) | 437.50<br>( 355.25,550.00 ) | 461.00<br>( 368.75,547.50 ) | 0.674 | 435.54±123.96 | 485.53±168.77 | 0.032 |
| LVEF ( % ) | 55.00( 45.00,59.00 ) | 51.00( 39.00,57.00 ) | 0.166 | 55.00( 47.25,59.00 ) | 52.00( 39.75,59.00 ) | 0.178 |
| IgCCL19<br>( pg/ml ) | 2.45±0.28 | 2.63±0.43 | 0.036 | 2.42 ( 2.28,2.60 ) | 2.45 ( 2.27,2.69 ) | 0.397 |
| IgCCL21<br>( pg/ml ) | 2.77 ( 2.58,2.91 ) | 2.95 ( 2.72,3.01 ) | 0.001 | 2.74 ( 2.57,2.90 ) | 2.88 ( 2.70,2.96 ) | 0.006 |
| IgNT-proBNP<br>( pg/ml ) | 3.49 ( 3.22,3.70 ) | 3.74 ( 3.50,4.10 ) | 0.001 | 3.41±0.42 | 3.64±0.40 | <0.001 |
| Medication |  |  |  |  |  |  |
| ACEI ( n,% ) | 5 ( 3.57 ) | 0 ( 0.00 ) | 0.588 | 4 ( 4.35 ) | 1 ( 1.28 ) | 0.238 |
| ARB ( n,% ) | 53 ( 37.86 ) | 6 ( 20.00 ) | 0.062 | 25 ( 32.05 ) | 34 ( 36.96 ) | 0.503 |
| BB ( n, % ) | 84 ( 60.00 ) | 14 ( 46.67 ) | 0.180 | 52 ( 56.52 ) | 46 ( 58.97 ) | 0.747 |
| CCB ( n, % ) | 35 ( 25.00 ) | 10 ( 33.33 ) | 0.477 | 19 ( 20.65 ) | 26 ( 33.33 ) | 0.062 |
| Diuretic<br>( n, % ) | 76 ( 54.29 ) | 16 ( 53.34 ) | 0.924 | 52 ( 56.52 ) | 40 ( 51.28 ) | 0.494 |
| Digoxin<br>( n, % ) | 31 ( 22.14 ) | 6 ( 20.00 ) | 0.989 | 16 ( 17.39 ) | 21 ( 26.92 ) | 0.133 |

SBP: systolic blood pressure; DBP: diastolic blood pressure; BMI: body mass index; TG: triglyceride; TC: total cholesterol; HDL-C: high density lipoprotein cholesterol; LDL-C: low density lipoprotein cholesterol; LVEF: left ventricular ejection fraction; ACEI: angiotensin-converting enzyme inhibitors; ARB: angiotensin receptor blockers. BB: $\beta$ -receptor blockers; CCB: calcium channel antagonist; NT-proBNP: N-terminal pro-B-type natriuretic peptide; CCL19: chemokine C-C motif ligand 19; CCL21: chemokine C-C motif ligand 21.

### Prognostic Value of CCL21

Analysis of the area under the curve (AUC) revealed that only CCL21 and NT-proBNP had statistically significant values. The AUC values for CCL19, CCL21 and NT-proBNP were 0.599 (95%CI, 0.481-0.717), 0.694 (95%CI, 0.589-0.800) and 0.694 (95%CI, 0.585-0.803), respectively, indicating their predictive ability for mortality. The optimal cut-off values for predicting all-cause mortality with the variables CCL19, CCL21 and NT-proBNP were determined to be 477.67 pg/mL, 831.76 pg/mL, and 4466.84 pg/mL, respectively. Furthermore, sensitivity and specificity analyses were conducted on these variables to predict all-cause mortality in heart failure patients individually. The sensitivity rates were found to be as follows: CCL21 60.0%, CCL19 36.7%, NT-proBNP 66.7%, The corresponding specificity rates were: CCL21 77.1%, CCL19 83.6%, NT-proBNP 68.6%. (*P*=0.089, *P*=0.001, *P*=0.001) (Figure 2A).

**Figure 2.**
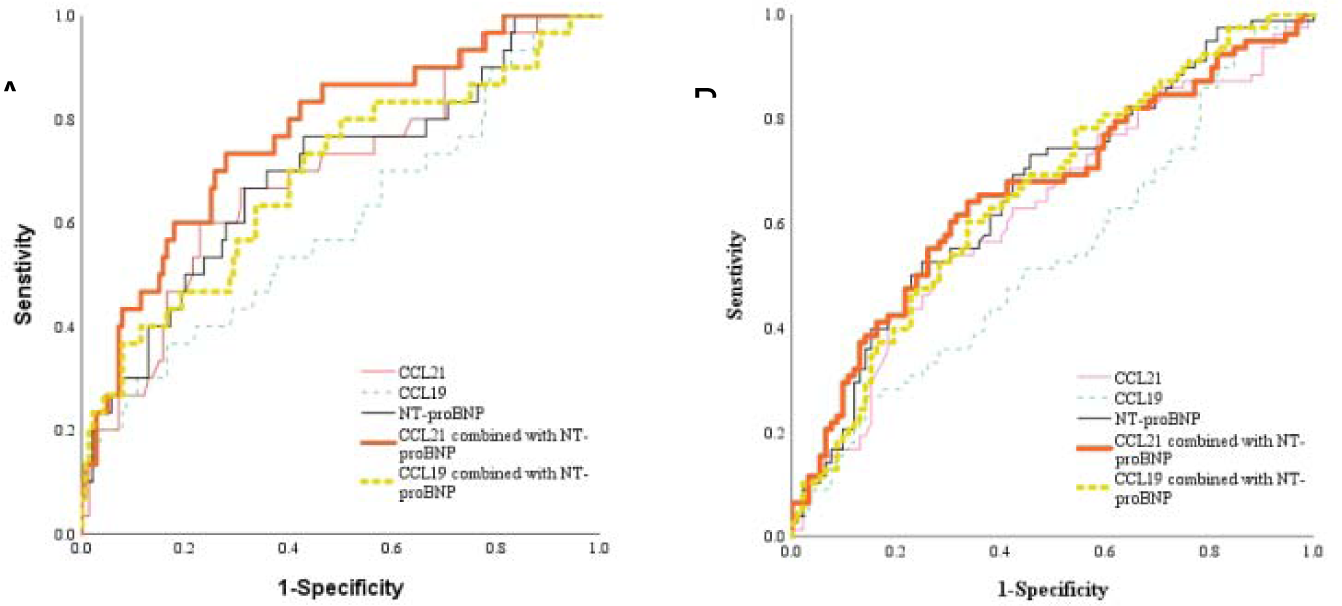
Receiver-operating characteristic curves of the predictive value of CCL19, CCL21, NT-proBNP and combining NT-proBNP for all-cause mortality(A) and composite endpoints(B) in patients with heart failure.

The analysis of AUC values revealed that only CCL21 and NT-proBNP showed statistically significant values in predicting composite endpoints. CCL19, on the other hand, was unable to predict the composite endpoints (the optimal cut-off value =478.63pg/ml, sensitivity=26.9%, specificity=85.9%, *P*=0.397), with an AUC was 0.538(95%CI, 0.450-0.625). The optimal cut-off value of CCL21 in predicting composite endpoints was found to be 741.31pg/ml, with a sensitivity of 52.6% and specificity of 72.5%. The corresponding AUC was calculated as 0.621 (95% CI, 0.536-0.706). For NT-proBNP, the best cut-off value was determined to be at 4466.84pg/ml, with a sensitivity of 52.6% and specificity of 75.0%. This yielded an AUC of 0.661(95%CI, 0.579-0.743). (Figure 2B).

More importantly, the value of combining CCL21 with NT-proBNP in terms of ROC prediction was analyzed. The AUC for predicting all-cause mortality was up to 0.769 (95%CI 0.675-0.863), with a sensitivity of 73.3% and specificity of 72.1%(*P*=0.001) (Figure 2A). For predicting the composite endpoints, the AUC was 0.662 (95%CI 0.597-0.760), with a sensitivity of 66.7% and specificity of 63.0%(*P*<0.001) (Figure 2B), These results indicate that the combing CCL21 with NT-proBNP improves the predictive effect for all-cause mortality and composite endpoints in heart failure patients.

The K-M method was proposed to assess the impact of different levels of CCL19, CCL21 and NT-proBNP on all-cause death and composite endpoints in heart failure patients. Due to the skewed distribution of NT-proBNP levels, they were logarithmically transformed to generate a linear trend and to minimize the effect of outliers. CCL19, CCL21 and NT-proBNP were categorized into reduction and augmentation groups using cut-off values of 477.67 pg/ml, 831.76 pg/ml and 4466.84 pg/ml, respectively. The results indicate that heart failure patients with elevated levels of CCL19, CCL21,and NT-proBNP have a higher risk of all-cause mortality (*P*<0.05,) (Figure 3).

**Figure 3.**
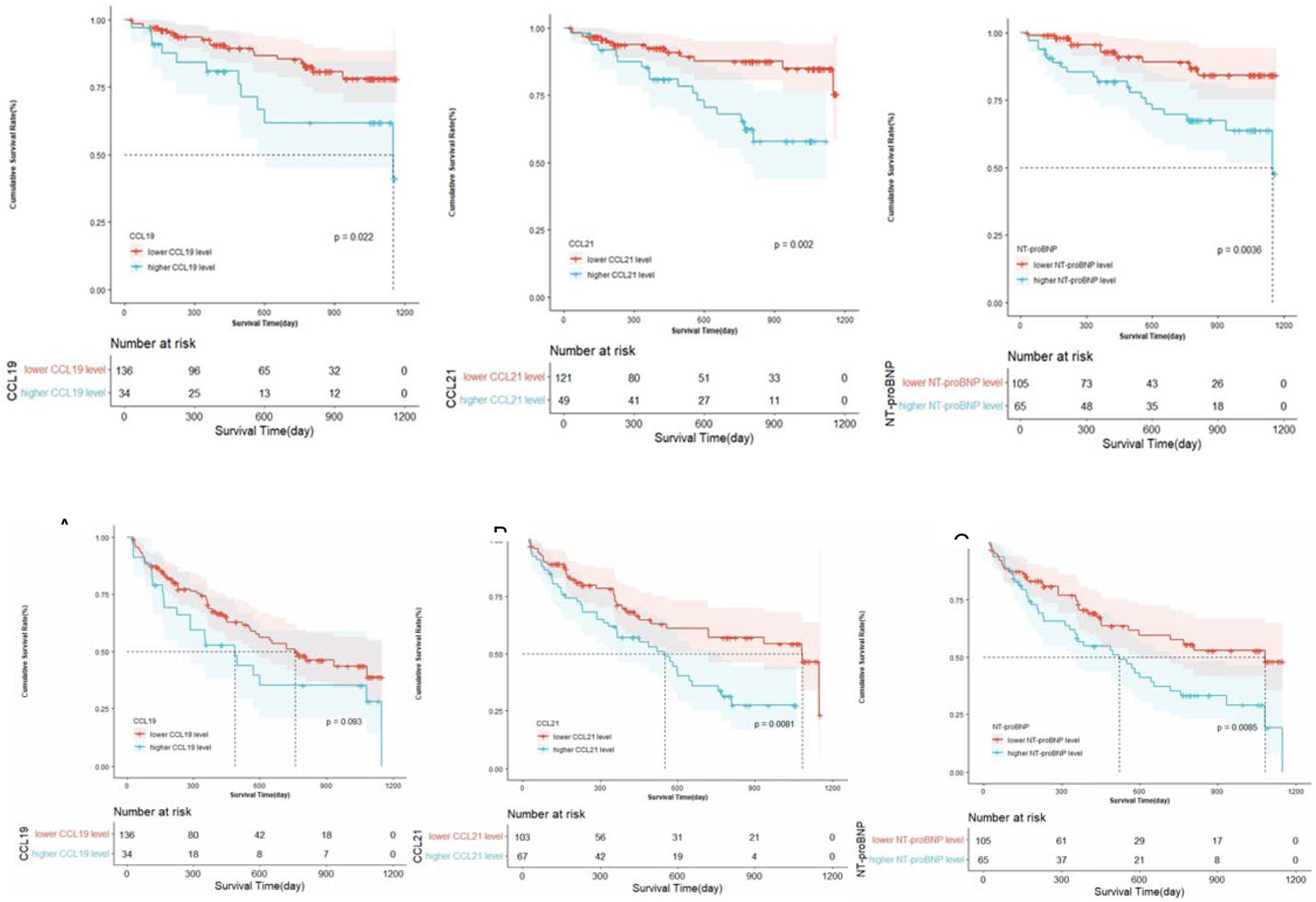
Kaplan-Meier curves analysis for the association of serum CCL19, CCL21, NT-proBNP level and incidence of all-cause mortalit, C) and composite endpoints (D, E, F).

Based on the ROC analysis of the optimal cutoff values for CCL19(478.63 pg/ml), CCL21(741.31 pg/ml), NT-proBNP (4466.84 pg/ml), heart failure patients were classified reduced groups and elevated groups. Results showed that heart failure patients with elevated CCL21(*P*<0.05) and NT-proBNP (*P*<0.05) had a higher risk of complex endpoint events. While no statistical significance was observed in CCL19(*P*> 0.05,) (Figure 3).

### Risk factors for heart failure

Univariate COX proportional hazard analysis revealed that elevated levels of CCL19 and CCL2, lgNT-proBNP, smoking, age, male, creatine and ischemic heart disease were associated with all-cause mortality(*P*<0.05), Table 2). Additionally, elevation in CCL21 levels, lgNT-proBNP levels, creatine levels, hypertension presence, diabetes presence, statins and CCBS use were found to be univariate correlation factors for complex endpoints (*P*<0.05, Table 2). The linear relationship between the above variables and mortality and complex endpoint events is analyzed without multivariate collinearity among the covariates. After adjusting for covariates, multivariate COX proportional hazard analysis revealed that smoking status, ischemic heart disease presence, elevated lgNT-proBNP levels and elevated CCL21 levels were associated with all-cause mortality (HRs= 2.585,2.342, 2.783, and 2.248, *P*=0.012, 0.027, 0.036, and 0.042, Table 3), only elevated CCL21 levels combined with diabetes presence were found to be associated with a composite endpoint in heart failure patients (HRs= 1.745 and 2.206, *P*=0.017 and 0.001, Table 3).

**Table 2.**
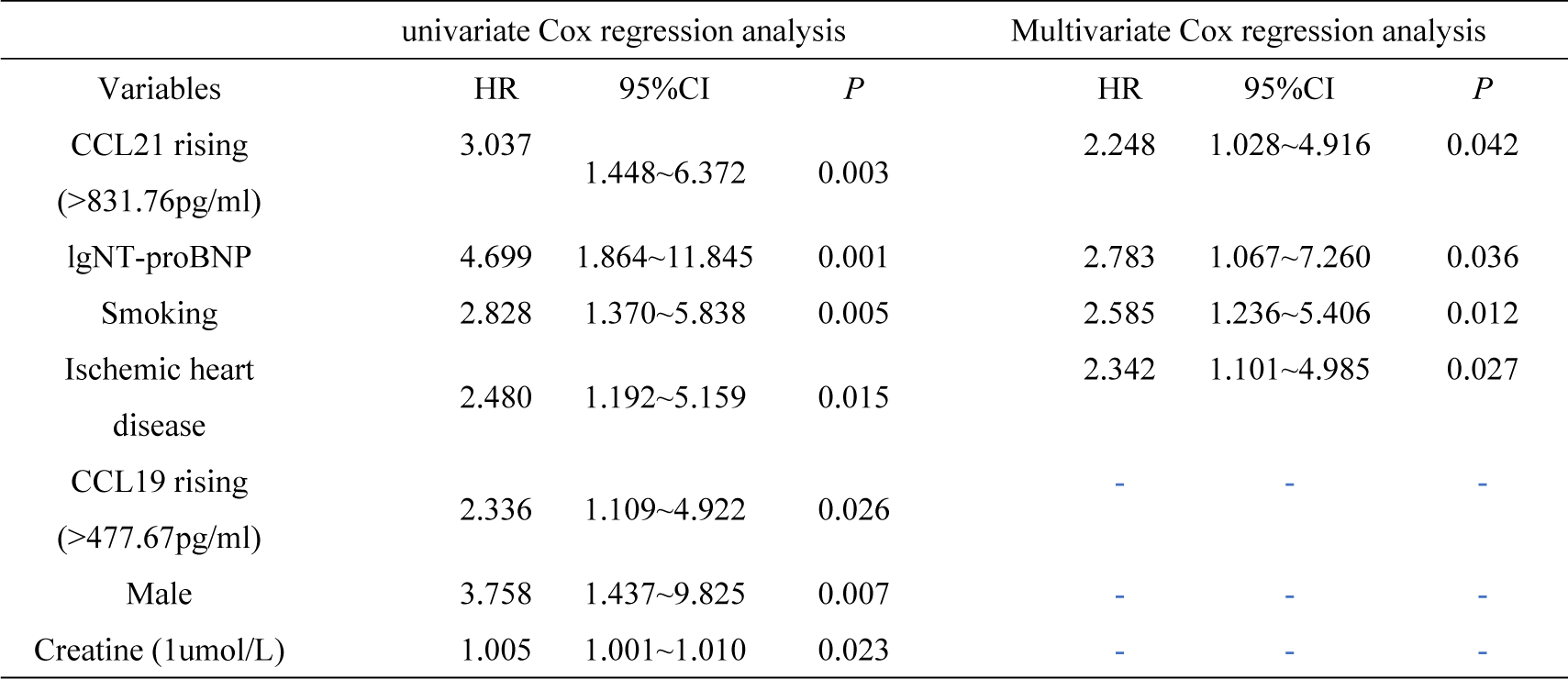
Univariable and Multivariable Analysis for all-cause mortality.

**Table 3.**
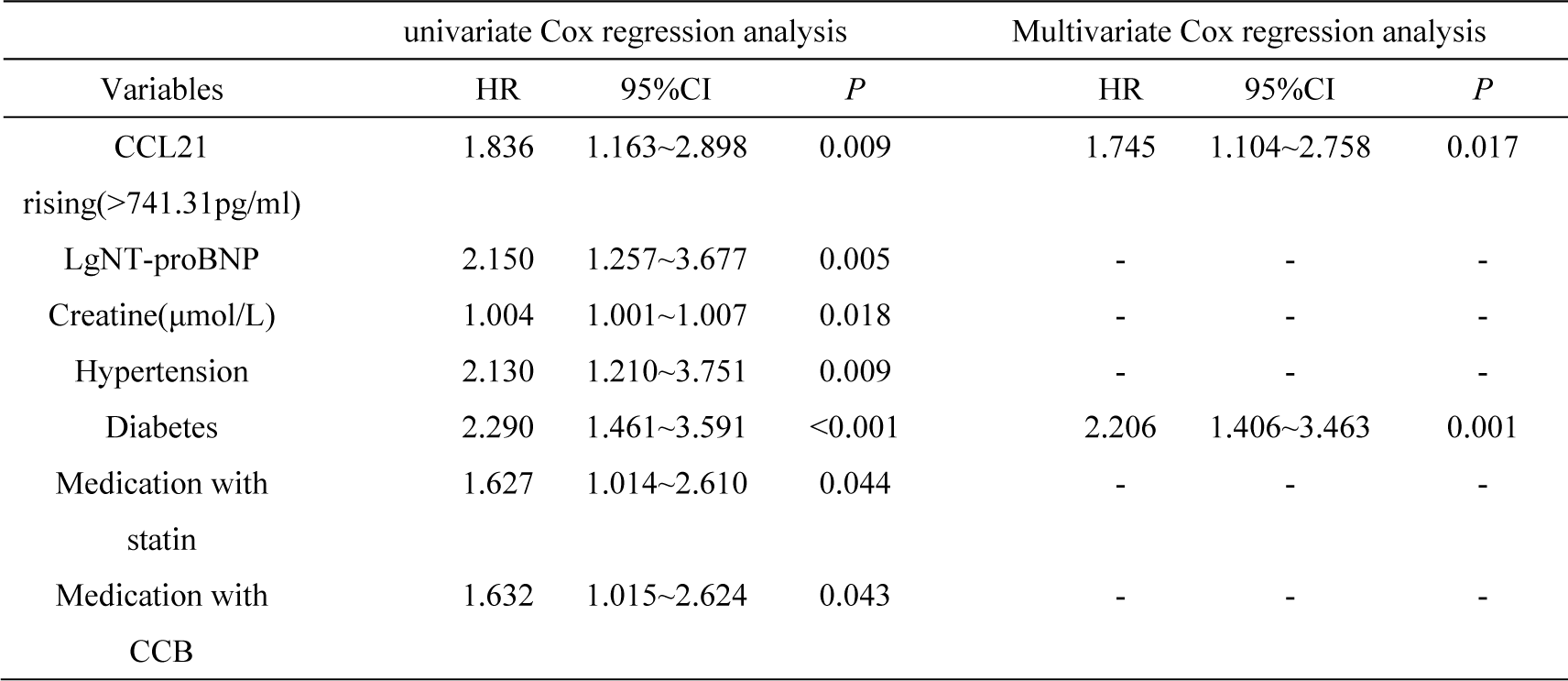
Univariable and Multivariable Analysis for composite endpoints.

### Correlation with Renal Function

Patients were divided into three groups based on their renal function: eGFR<30ml/(min×1.73m^2^) (G4/5), 30≤eGFR<60ml/(min×1.73m^2^) (G3), eGFR≥60ml/(min×1.73m^2^)(G1/2), The aim was to compare the levels of circulating biomarkers CCL19, CCL21 and NT-proBNP among patients with different renal insufficiency(Table 4). Significant statistical differences were observed in CCL 19 and NT-proBNP between different kidney function groups (*P*< 0.001), Serum CCL19 levels were higher in G3 and G4/5 stages compared to G1/2(*P*< 0.05), but no statistical significance difference was found in CCL21 level among the three groups (*P*> 0.05).

**Table 4.** Comparison of biomarkers in heart failure patients with different NYHA and Renal functions.

| Renal functions |  |  |  |
| --- | --- | --- | --- |
| G1/2<br>(n=99) | G3<br>(n=54) | G4/5<br>(n=17) | P |
| 2.40 ( 2.26,2.53 ) | 2.46 ( 2.26,2.66 ) a | 2.67<br>( 2.57,2.91 ) a | <0.001 |
| 2.78 ( 2.57,2.91 ) | 2.82 ( 2.64,2.96 ) | 2.87 | 0.284 |
|  |  | ( 2.64,2.97 ) |  |
| 3.44 ( 3.13,3.64 ) | 3.67 ( 3.27,3.85 ) a | 4.11 | <0.001 |
|  |  | ( 3.67,4.38 ) ab |  |
| a: means $P<0.05$ compared with the group of $\text{eGFR}\geq 60\text{ml}/(\text{min}\times 1.73\text{m}^2)$. b: means $P<0.05$ compared with the group of $30\leq \text{eGFR}<60\text{ml}/(\text{min}\times 1.73\text{m}^2)$. | | | |

Subsequently, our study demonstrated significant differences in CCL19 and NT-proBNP among groups with varying renal function (Pearson’s r=0.34, 0.41, both *P*<0.05), However, there were no significant differences in CCL21 between the three groups (Pearson’s r=0.12, *P*> 0.05), as depicted in Figure 4.

**Figure 4.**
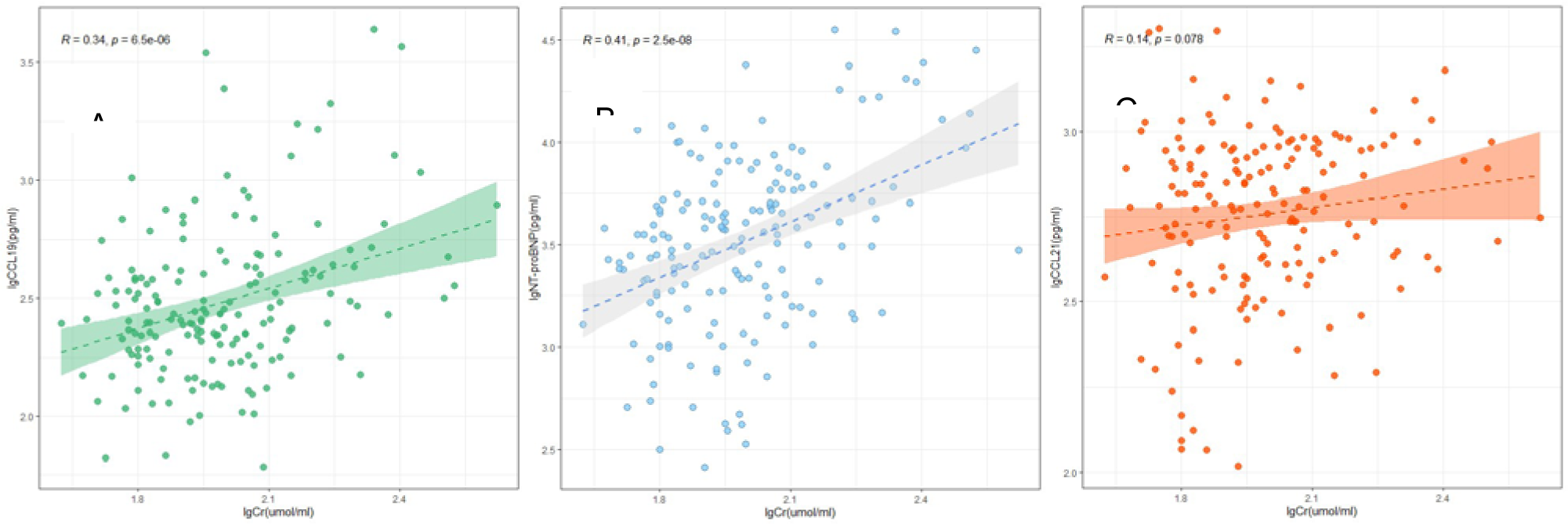
Correlation between the levels of CCL19(A), NT-proBNP(B), CCL21(C) and creatinine

## Discussion

The study found that significantly heart failure patients had significantly elevated serum concentrations of CCL21 and CCL19. CCL21 had strong predictive value for all-cause mortality and composite endpoints. This is the first report to show elevated serum levels of CCL21 in Chinese heart failure patients, which may be the result of fibrosis caused by inflammation of the heart, which in turn is triggered by recruitment of chemicals secreted by specific cell types in the heart [16,17]. Our main finding suggests that elevated serum levels of CCL21 are an independent risk factor for adverse prognosis in heart failure patients, even after adjusting the confounding factors. This discovery has the potential to improve predictions regarding mortality and adverse events related to heart failure. Furthermore, when combined with NT-proBNP for prediction, CCL21 offers additional value. While NT-proBNP is a powerful biomarker in heart failure, it does not encompass all the pathogenic processes of this complex disease. Therefore, relying solely on a single indicator cannot effectively reflect the risk stratification of heart failure patients. On the other hand, combining multiple markers’ circulating levels enhances the accuracy of disease prediction and holds greater clinical significance for at-risk patients [18].

The main risk factors for heart failure include coronary artery disease (CAD), hypertension, diabetes, family history of heart disease, obesity, etc. The ischemic heart disease is the leading cause of heart failure due to impaired perfusion of the heart muscles [1]. In this study, a majority of patients exhibited risk factors contributing to accelerated changes in the macroscopic structure of the heart. These changes includes reorganization and elongation of myocardial cells, resulting in ventricular hypertrophy and dilation, increased ventricular wall tension, and impaired subventricular perfusion. Microscopically, it is manifested by hypertrophy of cartilages, apoptosis and necrosis of cardiomyocytes, proliferation of fibroblasts, accumulation of proinflammatory mediators, and fibrosis. [19]. The immune system plays a critical role in the remodeling of the ventricles and contributes to both the inflammatory and repair phases. Studies have shown that GPR174/ CCR7 (CCL21 receptor) expression is elevated after myocardial injury, and activated CCL21-GPR174/ CCR7 signals are located on the myocardial fibroblasts of the injured myocardium [8]. In addition, not only did the study find that serum CCL21 levels in heart failure patients were associated with all-cause mortality, but also with composite endpoints including readmission due to heart failure, acute myocardial infarction and stroke. Nonetheless, NT-proBNP is susceptible to multiple cardiogenic and non-cardiogenic influences, with renal function having a greater impact on NT-proBNP. The PRIDE cohort study found that serum NT-proBNP levels were negatively correlated with renal function in both heart failure and non-heart failure patients, severely affecting the assessment of NT-proBNP in heart failure combined with renal insufficiency. It is difficult to determine whether elevated NT-proBNP levels are due to increased release due to cardiac disease or reduced clearance due to renal insufficiency [20]. However, after adjusting for confounding factors in this study, NT-proBNP was not associated with composite endpoints. On the contrary, the study found that CCL19 showed no association with mortality and adverse events in heart failure patients. The different association between CCL19 and CCL21 probably account for below. CCL19 is produced by various types of cells such as T cells, monocytes and macrophages, while CCL21 was primarily produced by stromal cell. What’s more, CCL21 has been found to induce more potent inflammatory effects in macrophages and have more powerful to chemotaxis than CCL19. In addition, CCL19 also integrates to atypical receptors CCX-CKR and CRAM to scavenge itself, which would influence the levels of CCL19 in patients [21,22]. Studies found that when cells were exposed to the same concentration gradient or the opposite gradient of CCL19 and CCL21 at the same time, the cells would preferentially migrate to the direction of CCL21, which may be related to the fact that the C-terminal of CCL21 itself contains 12 basic amino acid residues, which can bind closely with matrix proteoglycans and signals without internalization [23,24]. CCL19 can also bind to atypical chemokine receptors CCX-CKR (CCRL1) and CRAM (CCRL2) to endocytosis clear CCL19 and maintain the stable level of CCL19 in vivo [25], which may limit the continuous increase of serum CCL19 level to a certain extent and affect the expression of CCL19 in patients with severe disease. The study also found diabetes mellitus is an independent risk factor for complex terminal events in heart failure patients. Previous studies have found that the risk factors of heart failure in patients with diabetes are 2-4 times higher than those without diabetes, which is similar to the results of this study (HR=2.206) [26]. Studies have found that insulin resistance and hyperinsulinemia are associated with left ventricular hypertrophy and myocardial fibrosis [27]. Hyperglycemia and hyperinsulinemia can promote the activation of local renin-angiotensin-aldosterone system and the formation of advanced glycation products, aggravate the crosslinking of collagen molecules, cause increased myocardial stiffness, impaired myocardial diastolic function, and aggravate the degree of myocardial fibrosis [28,29]. While diabetes and heart failure are two diseases, they are separate and affect each other. Numerous clinical studies have found that patients with heart failure complicated with diabetes have worse prognosis [30,31]. The ESC heart failure Guidelines 2021 also states that heart failure patients with diabetes have a readmission rate 1.5 times higher than those without diabetes. It is suggested that strict control of heart failure comorbidity such as diabetes is helpful to reduce the readmitted frequency of heart failure patients and reduce the occurrence of complex endpoint events [1], which is also consistent with our findings.

The study suggested that the serum CCL21 could also be used as a prognostic biomarker for adverse events in patients with heart failure in Chinese people. These results were generally consistent with previous studies in western countries, Yndestad and Ueland found that serum levels of CCL21 were markedly raised in patients with heart failure, and associated with all-cause mortality independently [7,8]. Moreover, CCL21 was associated with all-cause and cardiovascular mortality [18], which was similar with our study. The same trend was seen in studies related to lung disease, where excessive levels of both chemicals were associated with poorer outcomes [32]. Lung diseases, such as pulmonary edema, are also present in heart failure patients, which also confirms the association between elevated levels of CCL21 and poor outcomes in heart failure patients.

Banas, B showed that CCR7 has a functional effect on the migration and proliferation of mesangial cells in the human kidney. SLC/CCL21 has a protective effect on mesangial cell survival [33]. Therefore, in this study, by comparing the changes in CCL21, CCL19, and NT-proBNP levels in patients with different levels of renal insufficiency, it was found that serum levels of CCL19 and NT-proBNP increased with renal function deterioration, while CCL21 levels were approximately unaffected by renal function. It has been suggested that CCL21 may be less affected by renal function and has the potential to be an influential complement to NT-proBNP, useful for clinicians to assess the status and long-term prognosis of heart failure patients. It partly explains the reason for CCL21 rather than NT-proBNP more linked with prognosis, because CCL21 rarely affected by poor renal function.

Study limitations acknowledged. Firstly, this was a single-center study and the population of our study was small. Secondly, heart failure patients were included preferred to have a serious condition, it is hard to expand the predictive value to the patients in stable state. Lastly, the study didn’t test the serum CCL19 and CCL21 continuously. It needs further study to exploit the relationship between changes of chemokines with prognosis of heart failure.

## Data Availability

The data used to support the findings of this study are included within the article.

## Notes

### Competing Interest Statement

The authors have declared no competing interest.

### Clinical Trial

K2019-05-032 K2021-01-031

### Funding Statement

This work was supported by the National Natural Science Foundation of China (No. 81873515), Natural Science Foundation of Fujian (No. 2020J011072), Medical Innovation Project of Fujian Province, China (Grant No. 2021CXA008) and high-level hospital foster grants from Fujian Provincial Hospital, Fujian province, China (No. 2020HSJJ03).

### Author Declarations

This study was conducted in accordance with the Helsinki Declaration and approved by the ethics committee of Fujian Provincial Hospital.

